# Microbial contributions to oxalate metabolism in health and disease

**DOI:** 10.1101/2020.01.27.20018770

**Authors:** Menghan Liu, Joseph C. Devlin, Jiyuan Hu, Angelina Volkova, Thomas W. Battaglia, Allyson Byrd, P’ng Loke, Huilin Li, Kelly V. Ruggles, Aristotelis Tsirigos, Martin J. Blaser, Lama Nazzal

## Abstract

Over-accumulation of oxalate in humans may lead to nephrolithiasis and nephrocalcinosis. Humans lack endogenous oxalate degradation pathways (ODP), but intestinal microbiota can degrade oxalate and protect against its absorption. However, the particular microbes that actively degrade oxalate *in vivo* are ill-defined, which restricts our ability to disentangle the underlying taxonomic contributions. Here we leverage large-scale multi-omics data (>3000 samples from >1000 subjects) to show that the human microbiota in health harbors diverse ODP-encoding microbial species, but an oxalate autotroph-*Oxalobacter formigenes-* dominates this function transcriptionally. Patients with Inflammatory Bowel Disease (IBD) are at significantly increased risk for disrupted oxalate homeostasis and calcium-oxalate nephrolithiasis. Here, by analyzing multi-omics data from the iHMP-IBD study, we demonstrate that the oxalate degradation function conferred by the intestinal microbiota is severely impaired in IBD patients. In parallel, the enteric oxalate levels of IBD patients are significantly elevated and associated with intestinal disease severity, which is consistent with the clinically known nephrolithiasis risk. The specific changes in ODP expression by several important taxa suggest that they play different roles in the IBD-induced nephrolithiasis risk.

## Main

Oxalate is a two-carbon molecule broadly present in nature [1]. In humans, oxalate from dietary sources can be absorbed through the gastrointestinal tract or produced endogenously from hepatic metabolism, and chiefly excreted into the urine. Lacking endogenous oxalate-metabolizing enzymes, all mammals are vulnerable to oxalate toxicity, which manifests as nephrolithiasis, nephrocalcinosis, chronic kidney disease (CKD), and rarely as life-threatening systemic oxalosis [2–5]. Nephrolithiasis affect 8% of the US population [6, 7] with an ∼20% 5-year recurrence rate [8, 9], and most kidney stones (85-90%) are composed of calcium oxalate[10]. Oxalate also has been implicated in inflammation and CKD progression, suggesting broad impact [11–14].

The mammalian intestinal microbiota can partially protect against oxalate toxicity by oxalate degradation [15–18]. Two recent large-scale epidemiological studies associated antibiotic use with increased nephrolithiasis risk [19, 20], presumably via a perturbed microbiota [21]. Multiple gut microbes can degrade oxalate [22], including *Oxalobacter formigenes*, an oxalate autotroph [23-27]. When given to animals, *O. formigenes* has reduced host oxalate burden [28–33]. *Escherichia coli, Bifidobacterium spp*. and *Lactobacillus spp*. also can degrade oxalate [34–43], but it is unclear which organisms are active oxalate degraders *in vivo*. A better understanding of human oxalate degrading microbes is needed to develop clinically protective strategies.

In particular, Inflammatory Bowel Disease (IBD) patients have heightened risk of oxalate toxicity due to elevated enteric oxalate levels and oxalate hyperabsorption, termed enteric hyperoxaluria (EH) [44–46]. Nephrolithiasis occurs in 12%-28% of adult IBD patients [44, 45]. Multiple factors are associated with EH in IBD patients, including lipid malabsorption and increased gut permeability. Another hypothesis is that microbiota-based oxalate degradation is impaired in IBD patients, leading to increased oxalate absorption [16].

Here, we leveraged large-scale multi-omics data to study oxalate degradation in the human microbiota. We curated all known microbial oxalate degradation pathways and identified those present in the human microbiota, and those that were transcriptionally active. We also interrogated the microbiota of IBD patients to understand shifts in microbiota-based oxalate degradation functions and their metabolic consequences.

## Results

### Type I and type II microbial oxalate degradation pathways (ODP)

To determine the ODP used by human gut bacteria, we curated all experimentally-validated microbial ODP from literature review and database searches (KEGG, MetaCyc, and Brenda) **(Fig. 1A)** [47-53]. There are two types of ODP based on their enzymatic mechanisms and co-factor requirements. Type I ODP cleave the oxalate carbon-carbon (C-C) bond in a single step **(Fig. 1A)** with no cofactor required. The two type I enzymes oxalate oxidase and oxalate decarboxylase **(Fig. 1A)** [54–57] are indistinguishable at the amino acid level [58] and belong to a single UniProt homologous protein family (IPR017774), therefore we refer to them jointly as oxalate oxidase/decarboxylase (OXDD). Type II ODP consists of two enzymatic reactions requiring coenzyme A as co-factor **(Fig. 1A)**. Initially, coenzyme A is added to oxalate to form oxalyl-CoA, e.g. by formyl-CoA transferase (FRC) **(Fig. 1A)**. In the second step, all type II ODP use oxalyl-CoA decarboxylase (OXC) to metabolize oxalyl-CoA into CO_2_ and formyl-CoA **(Fig. 1A)**.

**Figure 1.**
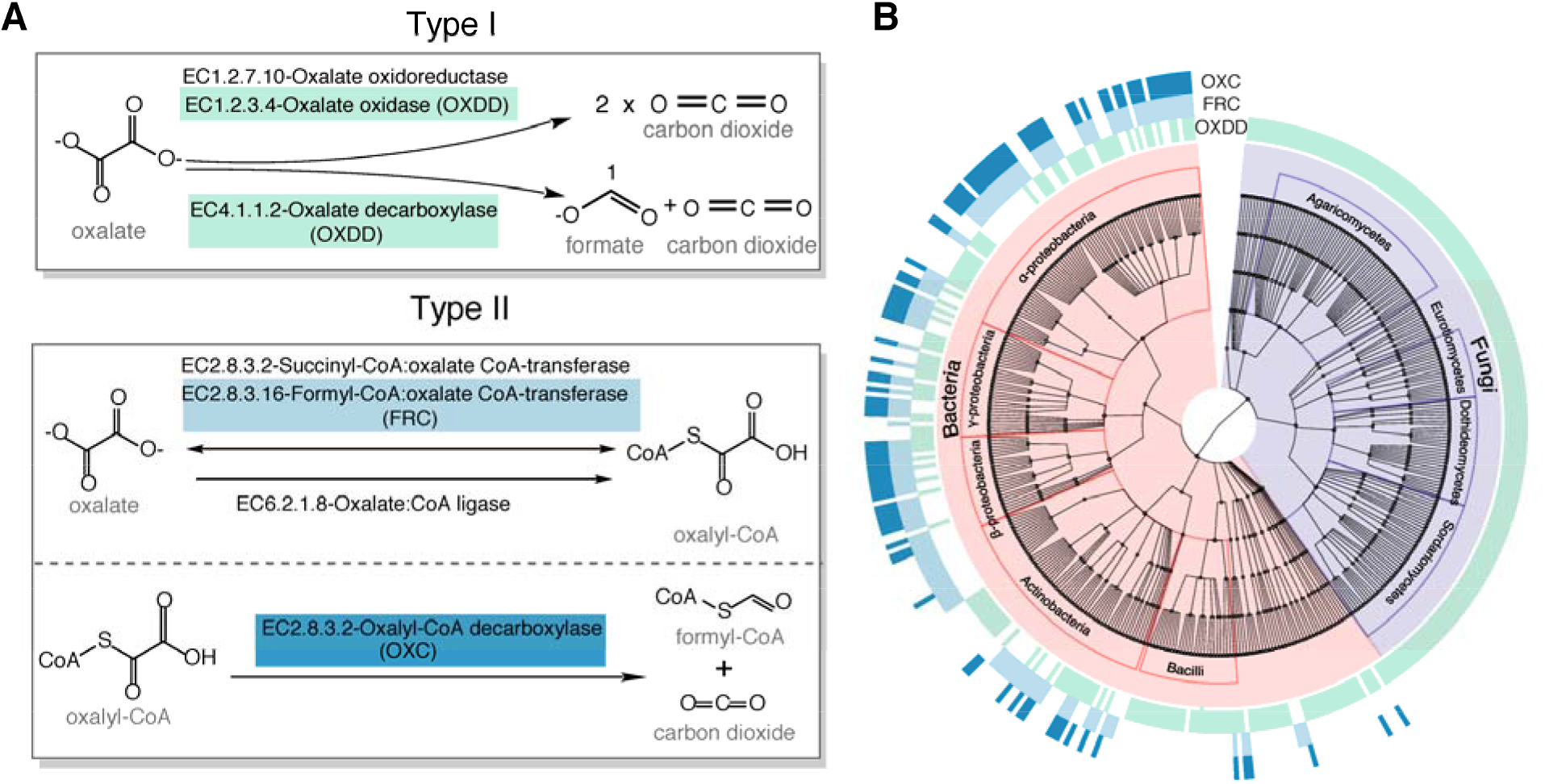
Type I and type II microbial oxalate-degrading pathway (ODP). **(A) Schema of type I and type II ODP**. Enzymes are annotated with corresponding KEGG IDs. OXDD, FRC, and OXC are the focus of the present study. **(B) Cladogram of microbial genera that encode oxalate-degrading enzymes OXDD, FRC, and OXC**. The three rings surrounding the cladogram indicate OXDD-, FRC- or OXC-encoding genera, respectively.

As such, we then acquired all available protein homologs of OXDD (n=2836), FRC (n=1947), and OXC (n=1284) from UniProt Interpro [59, 60]. By tracing the taxonomic origin of the genes encoding those homologs, we found that OXDD-coding taxa can be fungal or bacterial, whereas FRC- and OXC-coding taxa are strictly bacterial **(Fig. 1B)**. The frequent co-occurrence of FRC and OXC in individual genomes indicates encoding complete type II ODPs **(Fig. 1B)**. As expected, OXDD, FRC, and OXC each are conserved within the same microbial class, but exhibit substantial divergence across individual classes **(Fig. S1)**. These data provide a comprehensive overview of ODP pathways and a reference set of ODP-encoding microbes, which enable analyses to elucidate those relevant to humans.

### Type II ODP are utilized by the gut microbiota of healthy humans

Next, we ask which of these ODP are encoded and expressed within the metagenomes and metatranscriptomes of healthy humans **(Fig. S2)**. Multi-omics data from five studies were selected for downstream analyses, collectively including 2359 metagenome samples and 1053 metatranscriptome samples from 660 and 165 subjects, respectively **(Fig. S2, Table S1)**. After removing host-associated and low-quality reads, using DIAMOND Blastx [61] all remaining sequences were compared with the unique oxalate-degrading enzyme (ODE) homologous proteins, including 2519 OXDD, 1556 FRC, and 1037 OXC homologs. Alignment pairs with identity > 90% were retained for downstream analyses; this cut-off was determined by the protein identity of the inter- and intra-species ODEs to be robust for distinguishing ODEs that originated from different microbial species **(Fig. S3)**.

We found that ODEs were ubiquitously harbored in the healthy gut microbiome; at least one ODE was detected in the metagenome of 607 (92%) of the 660 subjects examined, and in the metatranscriptome of 132 (80%) of the 165 subjects available. In the metagenomes, the type II ODE FRC- and OXC-encoding genes *frc* and *oxc* were substantially more prevalent **(Fig. 2A)**, and more abundant **(Fig. 2B)** than the type I ODE OXDD-coding gene *oxdd*. Similarly, in the metatranscriptomes, *frc* and *oxc* expression also were substantially more prevalent **(Fig. 2C)**, and abundant **(Fig. 2D)** than *oxdd* expression. These findings were consistent in all studies, despite differences in source populations and sample preparation methods **(Table S1)**. In addition, specific *frc* and *oxc* genes **(Fig. S4)** were frequently co-expressed within the same microbiota, indicating the expression of the complete type II ODP. In total, these data indicate that the human microbiome is commonly and predominantly composed of microbes utilizing type II rather than type I ODP. As such, we focused on the type II ODP for the remaining analyzes.

**Figure 2.**
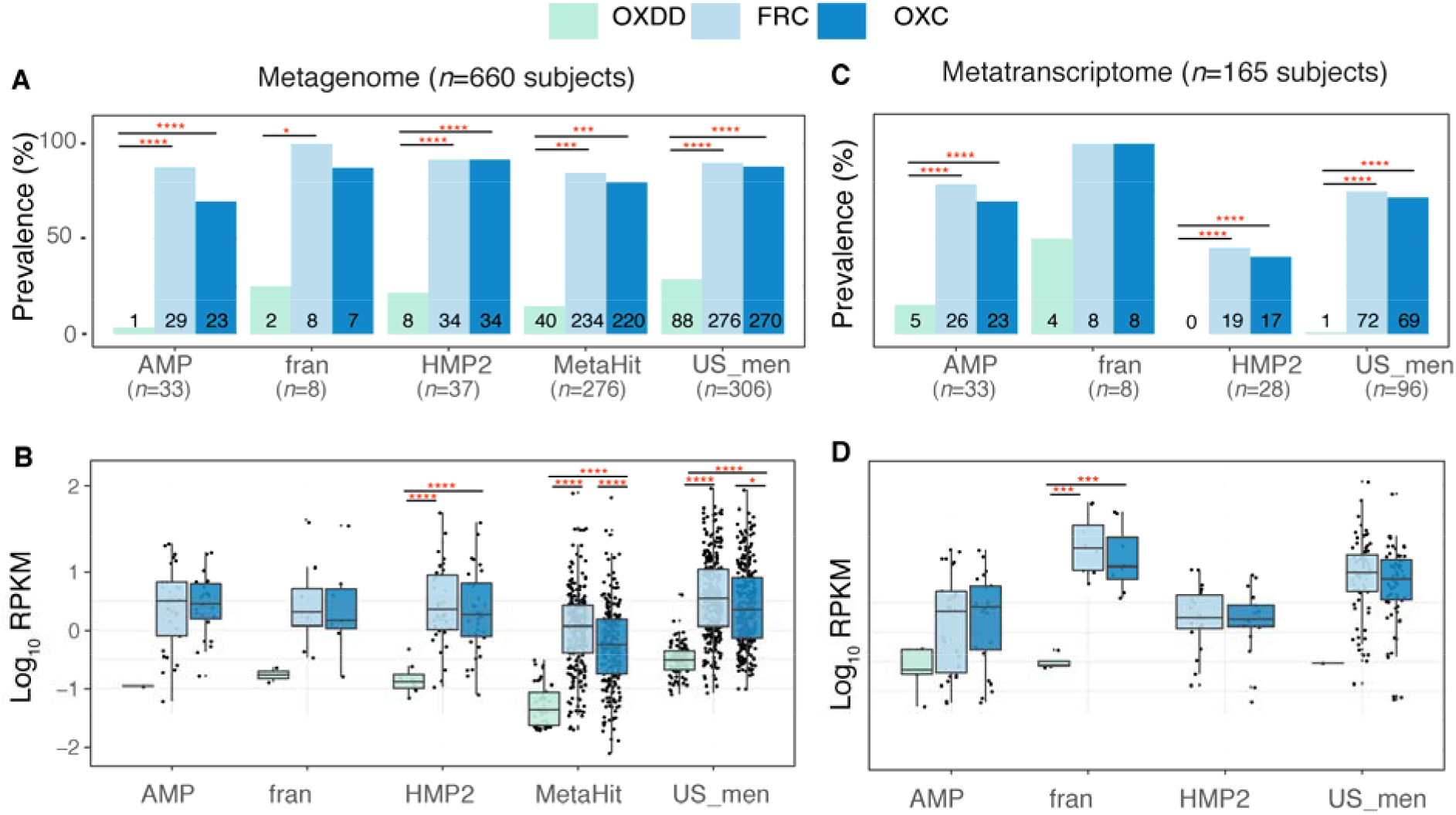
Detection of type I and II ODE within the fecal metagenome and metatranscriptome of 660 and 165 healthy human subjects. **Prevalence (A) and abundance (B) of ODE in the fecal metagenome of five studies surveyed**. Numbers written on the bottom bars indicate the numbers of subjects in whom the corresponding ODE is detected, and only those subjects were considered in panel B. **Prevalence (C) and abundance (D) of OXDD, FRC, and OXC in the fecal metatranscriptome of four studies surveyed**. *: p< 0.05, **: p< 0.01, ***: p< 0.001, ****: p< 0.0001, by proportion tests for panels A and C, by multiple-adjusted Mann-Whitney tests for panels B and C.

### ODPs are encoded by multiple microbes, but dominantly expressed by

#### *Oxalobacter formigenes* in healthy human microbiota

Although multiple microbes are known to encode *frc* and *oxc* [22, 62–64], their functional activity *in vivo* has not been studied. We next systematically characterized the active oxalate degraders *in vivo* by searching for the microbial species that transcribe these type II ODP genes. In the metagenome of 660 healthy subjects, we detected *oxc* encoded by multiple human gut microbes including *E. coli, O. formigenes, Muribaculaceae sp, and several Bifidobacterium sp*., *Lactobacillus sp*, **(Fig. 3A, left)**, some of which have been reported to degrade oxalate [31, 34–37, 41–43, 62, 65, 66]. The abundance of *oxc* of those species varied widely (median RPKM of 0.006-0.223) **(Fig. 3A, left)**. The most prevalent oxc-encoding microbe was *E. coli* (*oxc* detected in 56% of subjects), followed by *O. formigene*s (*oxc* detected in 39%). The abundance and prevalence pattern for *oxc* was consistent across different studies **(Fig. S5, S6)**, and was parallel to the other type II ODE *frc* **(Fig. S7)**,

**Figure 3.**
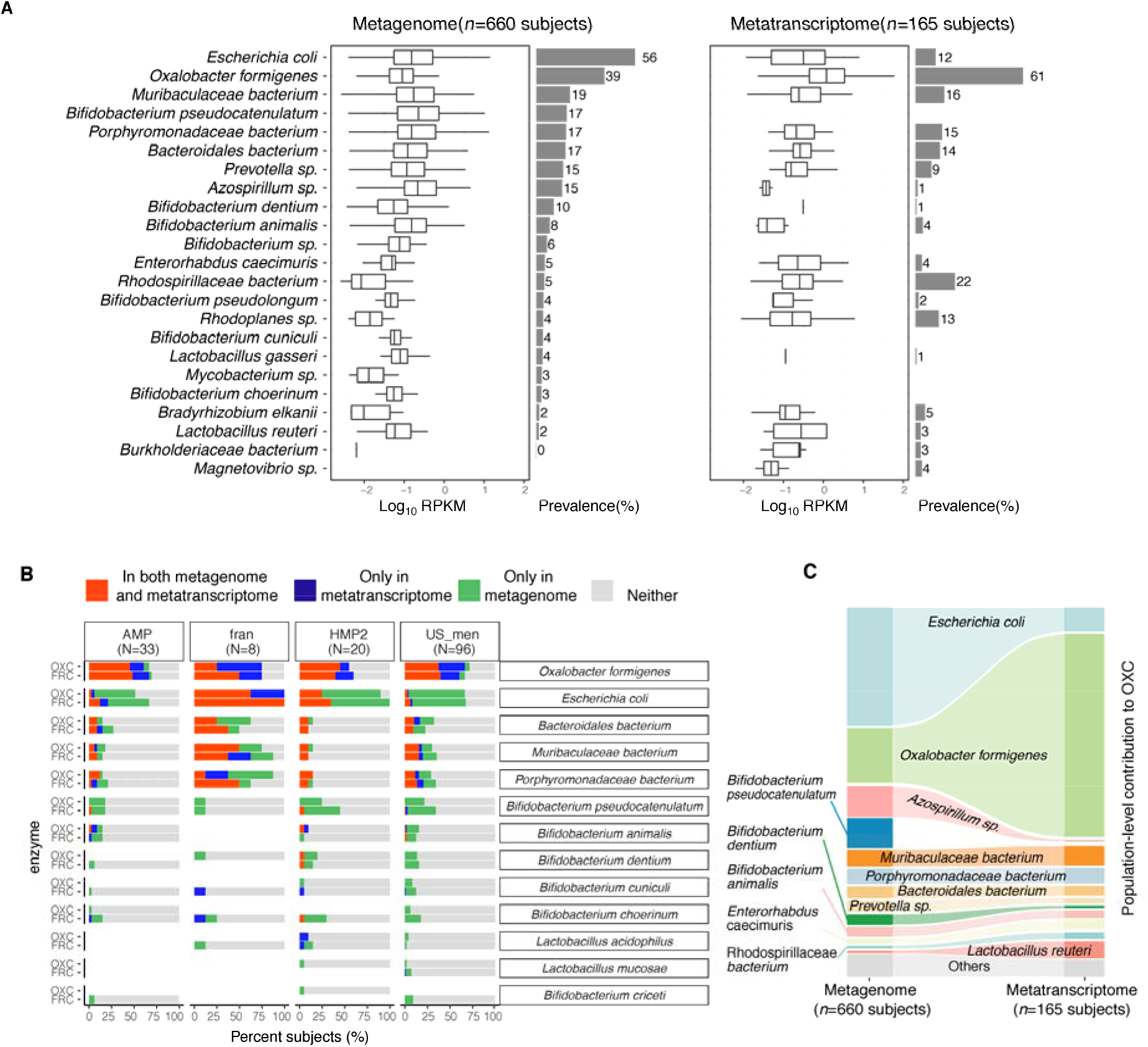
Expression of type II ODP of microbial species within the intestinal microbiota of healthy humans. **(A) Abundance and prevalence of OXC of microbial species in the metagenome (left) or metatranscriptome (right) of 660 and 165 subjects**. Box plots indicate the abundance of microbial OXC (log_10_ RPKM) among subjects in whom OXC is detected, and are generated with *ggplot2* with outliers excluded. Bar plots indicate the prevalence of microbial oxc, with percentage annotated. Microbial species are ordered by the corresponding metagenomic OXC prevalence. A parallel analysis for FRC is shown in Suppl. Fig 5. **(B) Detection of OXC and FRC of microbial species in the subject-matched metagenome and metatranscriptome, by study**. For each microbial ODE, the subjects are divided into four groups (shown in different colors) based on the co-detection of ODE in the matched metagenome and metatranscriptome, with percent (%) of which reflected. The fran Study, from which *E. coli* ODP was detected in all subjects, used a sample extraction method known to induce *E. coli*, as noted in their publication [92]. **(C) Population-level contribution of individual species to metagenomic (left) or metatranscriptomic (right) OXC**. The population level-contribution of each species was calculated at a relative scale (see Methods) and plotted. Raw values can be found in Suppl. Table 1. The 10 species that have the highest metagenomic or metatranscriptomic contribution are shown. A parallel analysis for FRC is shown in Suppl. Fig. 7.

Surprisingly, *oxc* expression across species did not directly correlate with *oxc* metagenomic abundance or prevalence **(Fig. 3A, right)**. For *O. formigenes*, whose growth relies on oxalate degradation [23, 67, 68], *oxc* expression was both most abundant and most prevalent, present in the metatranscriptome of 61% of subjects **(Fig. 3A, right)**. In contrast, *E. coli oxc* expression was observed in only 12% of subjects **(Fig. 3A, right)**, despite gene presence in 56% of subjects **(Fig. 3A, left)**. For *Bifidobacterium* and *Lactobacillus* species, whose oxalate degradation activity was reported *in vitro* and in animal models [42, 43, 66], *oxc* expression was observed at low prevalence (< 5%), or not observed at all (*B. pseudocatenulatum, B. choerinum*) **(Fig. 3A right)**. Collectively, these data demonstrate variation in the extent of ODP transcription by species and individual hosts.

To test the species-specific expression pattern of ODP in a more rigorous manner, we restricted our analysis to the subject-matched metagenome and metatranscriptome that derived from the same microbiota communities **(Fig. 3B)**. *O. formigenes* ODP expression was observed in most subjects whose microbiome harbored the corresponding gene **(Fig. 3B)**, and in many subjects in whom the gene was not detected, indicating underdectection of *O. formigenes* ODP at the genomic level. In contrast, *E. coli* ODP expression was only observed in a small portion of subjects harboring the corresponding genes **(Fig. 3B)**. The relative lack of detection of *O. formigenes* ODP genes in the metagenomic data may reflect the highly variable abundance of the organism itself falling at times below the lower limit of detection using genetic-based methods [69–72].

We then assessed population-level contribution (see Methods) to quantify the impact of individual species on global ODP. The contribution of *O. formigenes* to ODP increased from 17% to 63% from the metagenomic to the metatranscriptomic level, greater than the transcriptomic contributions of all other species combined **(Fig. 3C, Table S2)**. Conversely, the contribution of *E. coli* to ODP was markedly reduced from the metagenomic (36%) to the metatranscriptomic (7%) level **(Fig. 3C)**. The contributions of other species to ODP at the metagenomic and metatranscriptomic levels varied, but none were dominant **(Fig. 3C)**. A parallel pattern was observed for *frc* **(Fig. S8, Table S3)**. Network analysis did not yield significant species-species interactions related to *oxc* transcription, potentially because the activity of non-*O. formigenes* species was too low across the studies (data not shown).

In summary, we systematically described the gut microbes responsible for oxalate degradation *in vivo* in health, showing that this microbial function is dominated by a single species-*O. formigenes*. These data provide a baseline for examining the disease-associated changes in subsequent analyses.

### Impaired microbiota-based oxalate degradation and elevated enteric oxalate levels in IBD patients

IBD patients frequently suffer from Enteric Hyperoxaluria (EH), with oxalate hyperabsorption and formation of calcium oxalate kidney stones [44–46]. In particular, Crohn’s disease (CD) with ileocolonic involvement is associated with greater nephrolithiasis risk compared to either ileal or colonic involvement alone [73]; Ulcerative colitis (UC), regardless of severity and location, is associated with stone formation [73]. We hypothesized that the oxalate degradation function conferred by the intestinal microbiome may be impaired in IBD patients, ultimately leading to more intestinal and circulating oxalate in the host. To address this question, we acquired the multi-omics iHMP-IBD study data [74, 75] to assess both oxalate homeostasis and microbiota-m based oxalate degradation.

We stratified the iHMP-IBD patients into UC (N=30), CD (N=54) groups, regardless of the disease location, and a CD subgroup with the ileocolonic phenotype at baseline - CD-L3 (n=25). Consistent with the clinical nephrolithiasis risk, fecal oxalate concentrations were elevated in both the UC patients (*p*=0.005) and CD-L3 patients (*p*<0.001) compared to healthy controls; the same trend was observed in the CD patients (p=0.06) **(Fig. 4A)**. Fecal oxalate was most increased in the CD-L3 patients **(Fig. 4A, Fig. S9)**. We also examined the differences in fecal oxalate concentrations in a linear mixed-effects model, adjusted for subject age, and sex as fixed effects and repeated measurements as a random effect, and observed the effects at marginal levels (*p*=0.095 for CD, *p*=0.083 for UC, *p*=0.058 for CD-L3).

**Figure 4.**
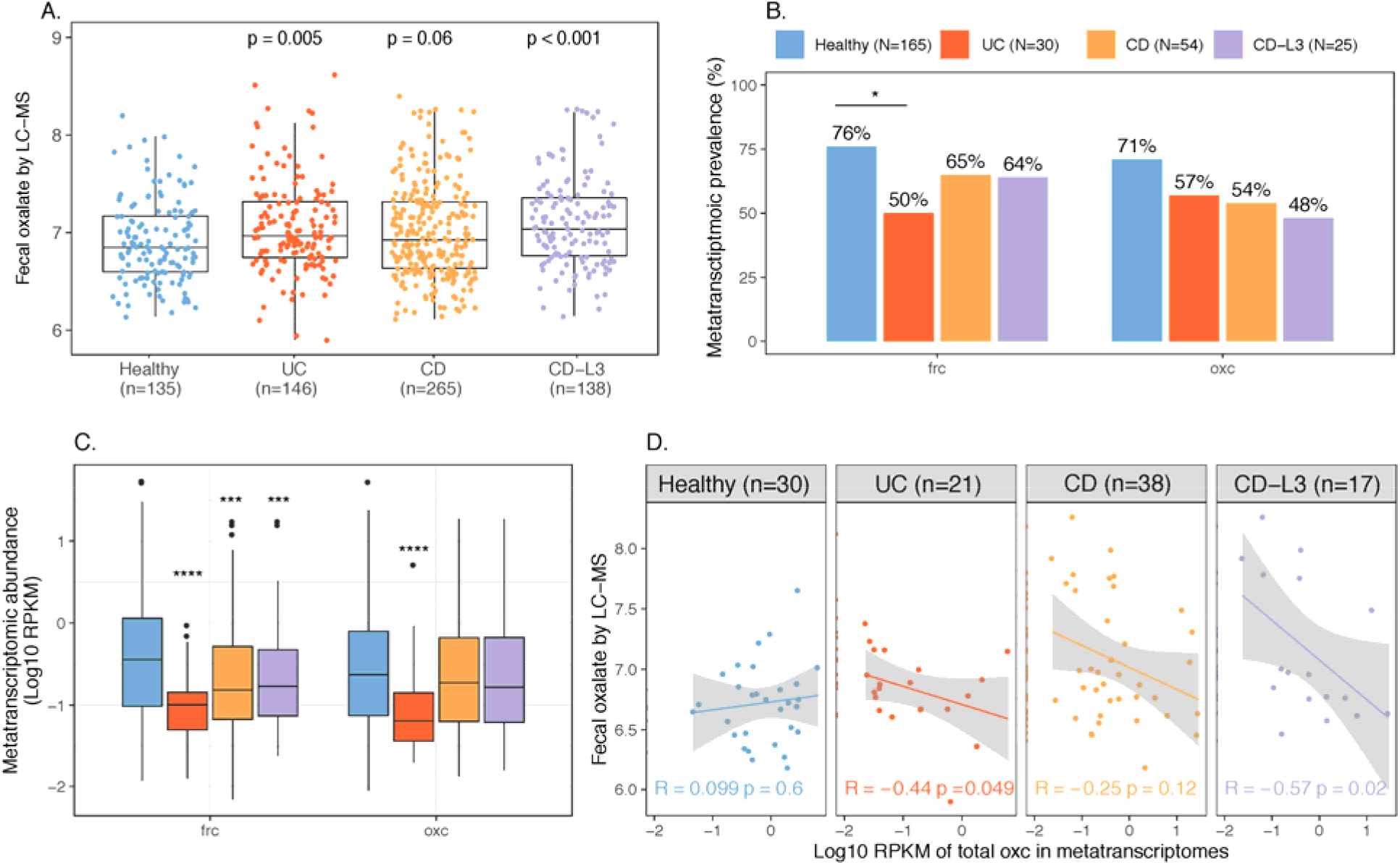
Elevated fecal oxalate concentrations and reduced expression of microbiome ODP in IBD patients. **(A) Stool oxalate concentrations in healthy, UC, CD, or CD-L3 subjects from HMP-IBD study**. L3 refers to the ileocolonic phenotype, according to the Montreal Classification) at baseline. Data derived from iHMP-IBD untargeted metabolomics measurements. **Prevalence (B) and abundance (C) of OXDD, FRC, and OXC in metatranscriptomes of healthy, UC, CD, or CD-L3 subjects**. The 165 healthy controls are combined from four studies (AMP, US_men, fran, HMP2). *: p< 0.05, **: p< 0.01, ***: p< 0.001, ****: p< 0.0001 by multiple-adjusted Mann-Whitney tests in A and C, by proportion test in B. (D). Spearman correlations of fecal oxalate and total metatranscriptomic OXC. Spearman Rho and p values are shown.

Expression of *frc* in the gut microbiota was only observed in 50% of UC (*p*<0.05), 65% of CD and 64% of CD-L3 patients compared to 76% in healthy subjects **(Fig. 4B)**, and the levels of expression were significantly decreased in all IBD groups (*p*<0.001) **(Fig. 4C)**. Similarly, *oxc* expression was observed in 57% UC, 54% CD and 48% CD-L3 patients **(Fig. 4B)** compared to 71% in healthy persons, and was significantly downregulated in the UC group (*p*<0.001) **(Fig. 4C)**. Type I ODE *oxdd* was rarely observed in the IBD microbiota, similar to healthy persons (data not shown).

We then asked whether there was a relationship between the decreased microbiota-based ODP expression and oxalate levels in the intestinal tract. By Spearman correlation, 44%-57% of the variance in fecal oxalate could be explained by variation in ODP expression in IBD patients (p<0.05 for UC and CD-L3 subjects) **(Fig 4D)**. In contrast, microbial ODP genes showed increased abundance in IBD patients **(Fig. S10 A, B)**, indicating the discrepancies between the gene abundance and expression levels.

Collectively, these data indicate that IBD patients have decreased microbiota-based oxalate degradation; the observed elevated intestinal oxalate may be a consequence, conferring increased susceptibility to EH.

### Loss of *O. formigenes* and downregulation of its ODP in IBD patients

We next sought to identify which microbial species accounted for the reduced ODP expression in IBD patients. The two species *E. coli* and *O. formigenes* with the largest ODP contributions at the genomic and transcriptional level, respectively, were notable.

Using gene and transcript jointly as markers, ODP expression by *O. formigenes* was detected in only ∼25% in UC and in CD patients **(Fig. 5A)** - less prevalent than the ∼70% in healthy individuals, either when studies were combined **(Fig. 5A)** or separate **(Fig. 3D)**. Having the lowest *O. formigenes* prevalence, the CD-L3 group had strongest negative correlation between fecal oxalate and expression of *oxc* **(Fig. 4D)**. Consistent with the low prevalence, the ODP expression by *O. formigenes* was also less abundant in UC, CD, and in CD-L3 patients compared with controls (p<0.01 for all groups), marked by lower RPKM values **(Fig. 5B)**, Interestingly, *O. formigenes* ODP was always actively expressed, when genes were observed, within the microbiota of the IBD subjects **(Fig. 5A)**. In contrast, the ODP of *E. coli* was detected in nearly all IBD subjects and was transcribed more frequently compared to healthy subjects **(Fig. 5A)**.

**Figure 5.**
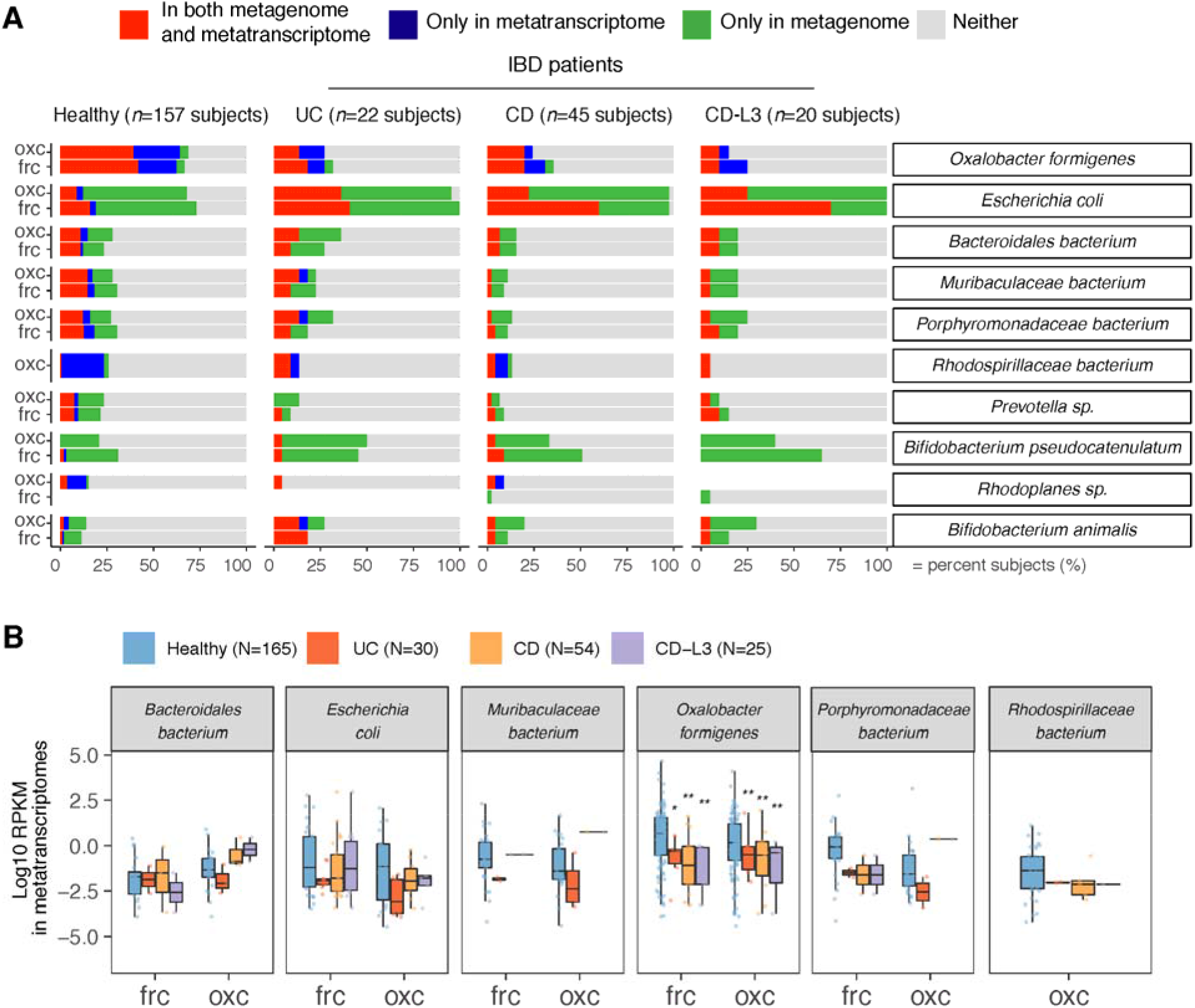
Differential ODP expression by human gut microbes in healthy and disease states. **(A) Detection of microbial OXC and FRC in the subject-matched metagenome and metatranscriptome from healthy subjects, UC, CD, or CD-L3 patients**. For each species shown, the subjects are divided into one of four categories based on the co-detection of ODE in the matched metagenome and metatranscriptome. **(B) Expression of microbial FRC and OXC in the metatranscriptomes of healthy subjects, UC, CD, or CD-L3 patients**. Boxplot reflects the subjects, in whose metatranscriptome the corresponding enzyme is detected. * p <0.01, ** <0.0001 by multiple-adjusted Mann-Whitney tests.

In total, these data revealed the complex pattern of taxa-specific ODP expression change during a disease state. In particular, the contrasting patterns of *O. formigenes* and *E. coli* illustrate the varied responses in association with distinct disease progression. Abolishing the ODP expression of *O. formigenes*, leads to the impaired global oxalate degradation, which further leads to increased circulating oxalate. Although *E. coli, Lactobacillus* spp, and *Bifidobacterium* spp, use their ODPs to defend against oxalate-induced acid stress [34, 41, 62, 64, 66], their upregulated ODP in IBD is a secondary response to the elevated oxalate levels present.

## Discussion

Oxalate degradation by the human microbiota has been known since the 1940s [15, 17, 18, 49, 68, 76, 77], but the taxa involved *in vivo* were not clear [78, 79]. We present the first comprehensive study of human oxalate-degrading microbes, with multi-omics evidence, and quantitatively characterized their contributions. Our finding that multiple human gut microbes encode ODPs is consistent with multiple prior studies [22, 43, 64, 80, 81], but surprisingly, oxalate degradation in the gut microbiome was dominated by a single organism - *O. formigenes*, which is the only known oxalate-degrading specialist that exclusively uses oxalate as energy and growth sources [22]. By actively transcribing ODPs, *O. formigenes* dominates the global ODP of the microbiota, consistent with a previous proteomic analysis of *O. formigenes* cells, showing FRC and OXC as the most abundant proteins produced during exponential and lag stages [82].

Most kidney stones (85-90%) are idiopathic with few genetic associations [83]. Here using the risk population for nephrolithiasis – IBD patients as the surrogate, we provide novel insights that the impaired metabolic activity of the microbiota may mediate enteric oxalate levels and nephrolithiasis risk. We showed that fecal oxalate concentrations were elevated in the iHMP-IBD UC patients, and CD patients with ileocolonic involvement, which is consistent with the clinical observations of high EH risk [73, 84, 85]. EH may be an attractive indication for *microbiota-*associated therapy, as the elevated soluble oxalate is in the intestine, providing an advantage for oxalate degrading bacteria, such as *O. formigenes* to colonize and to be metabolically active. Our finding that *O. formigenes* is less common in IBD patients than in healthy individuals provides a rationale for restoring *O. formigenes* in this population. Previously *O. formigenes*-related therapeutics was only examined in patients with primary hyperoxaluria (PH) and yielded inconsistent results [28, 29, 86, 87]. In contrast to EH, PH is caused by high circulating oxalate of host origin, therefore *O. formigenes*-based therapy may be limited by oxalate secretion into the intestinal lumen [31, 88, 89] for direct bacterial access.

The contrasting genetic and transcriptional changes in ODP observed in this study highlight the importance of evidence beyond the gene level for microbiome studies. The integrative multi-omics analysis framework built for this study, now deposited on Github, can also be extended to a broad range of microbiome functions of interest, such as choline and tryptophan metabolism, for example.

## Methods

### Meta-omics data of the human microbiome

Metagenomic and metatranscriptomic data of healthy human subjects were collected from 5 studies and 4 studies, respectively[75, 90-95]. Metagenomic and metatranscriptomic data of healthy humans and IBD subjects were collected from the iHMP-IBD study [75]. Each sample was cleaned by KneaData to remove low-quality reads and host-associated reads. The metabolic profiles of each sample were surveyed using HUMAnN2 v0.11.1 [96] under parameters --prescreen-threshold 0.01, --pathways-database metacyc_reactions_level4, metacyc_pathways_structured and --protein-database uniref50, for the comparison in **Fig. S2**. Fecal oxalate concentration was determined from untargeted metabolomics data from iHMP-IBD; measurements related to oxalate were selected for analysis.

### Homologous proteins of ODE

The homologs protein families of OXDD, FRC, and OXC were characterized by UniProt Interpro [59, 60] (V70) in protein families IPR017774, IPR017659, and IPR017660, respectively. We acquired the taxonomic origin and amino acid sequences of 2,699 OXDD, 1,947 FRC, and 1,284 OXC homologs. Protein homologs that are 100% identical were then de-duplicated into 2519 OXDD, 1556 FRC, and 1037 OXC unique homologs, which were used as a reference database of ODEs for a subsequent query against the meta’omics data. Oxalate oxidoreductase **(Fig. 1A)**, a recently discovered enzyme with limited information [51, 52, 97–101], was not considered in this present study.

### Pairwise identity between ODE protein homologs

Multiple sequence alignments were performed among the 2519 OXDD, 1556 FRC, and 1037 OXC unique protein homologs separately, by muscle [102] in seaview v4.7 [103]. The alignments then were trimmed and imported into R. The pairwise alignment distance d was calculated using function *dist*.*alignment* in the seqinR package [104] based on identity or Fitch matrix [105]. The alignment distance d was subsequently converted to percent protein identity 100 * (1 - *d*^2^), following the documentation of *dist*.*alignment*.

### Detection of ODE in the meta’omics data

The quality-filtered meta’omics data were aligned against the reference protein databases consisting of 2519 OXDD, 1556 FRC, and 1037 OXC unique homologs, by diamond blastx [61], with best hit returned (--max-target-seqs 1). Alignments with identity > 90% were kept for downstream analysis to achieve species-level resolution for most taxa **(Fig. S3)** The abundance of each ODE protein homolog was calculated as reads per kilobase per million (RPKM) in each sample. When multiple timepoints were available, each subject was represented by the mean measurements across all samples provided.

### Population-level contribution to ODE

The population-level contribution of a species to ODE was designed as a measurement to take both prevalence and abundance information into consideration. It is calculated for each ODE separately, based on their abundances (RPKM values).

Using oxc as the example, suppose there are *M oxc*-coding species and *N* samples. In any given sample *j*, the contribution of species *i* to OXC, *c*_*ij*_, is represented by its relative oxc abundance, calculated from

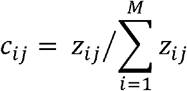

where the *z*_*ij*_ denotes the RPKM_oxc_ of species *i* in sample *j*. In this way, we normalize across samples with the total contribution in any OXC-positive samples is fixed to 1, in any OXC-negative samples is 0.

The population-level contribution of species *i* : *C*_*i*_, can be subsequently calculated from summating contribution of species *i* in *N* samples, as follows

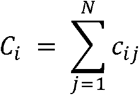

Note that population-level contribution of species monotonically increases with sample size *N*.

Therefore, it is transformed to relative scale when being compared across different populations or different sample types (metagenome vs. metatranscriptome), such as in **Fig. 3C, Fig. S8**, and **12**.

### Network analysis

A network analysis on *oxc* or *frc* expression from microbial species using SpiecEasi[106]. The raw RPKM values were used and the networks were constructed under default parameter *method=‘mb’, sel*.*criterion=‘bstars’, lambda*.*min*.*ratio=2e-2,nlambda=100, pulsar*.*params=list(rep*.*num=20, ncores=2)*.

## Data Availability

Source code of the pipeline can be found on Github via https://github.com/ml3958/FindTaxaCtrbt. Downstream analyses scripts are available per request.

## Acknowledgements

We thank Dr. David Goldfarb, Dr. David Fenyo, Dr. Victor Torres, Dr. Joao Xavier and Xuhui Zheng for their helpful comments. This study was supported in part by U01AI22285, R01DK110014, and the Rare Kidney Stone Consortium (U54 DK083908) from the National Institutes of Health, by the C & D and Zlinkoff Funds, Oxalosis and Hyperoxaluria Foundation-American Society of Nephrology career development grant, and the TransAtlantic Partnership of the Fondation LeDucq.

## Contributions

M.L, L.N, and M.J.B designed the study and wrote the manuscript. M.L, J.C.D, J.H performed the analyses. M.L, J.C.D, A.V, T.B collected the data. A.B, P.L, H.L, K.V.R, A.T contributed by suggesting additional new analysis and associated methods.

## Supplementary Figures

**Figure S1.**
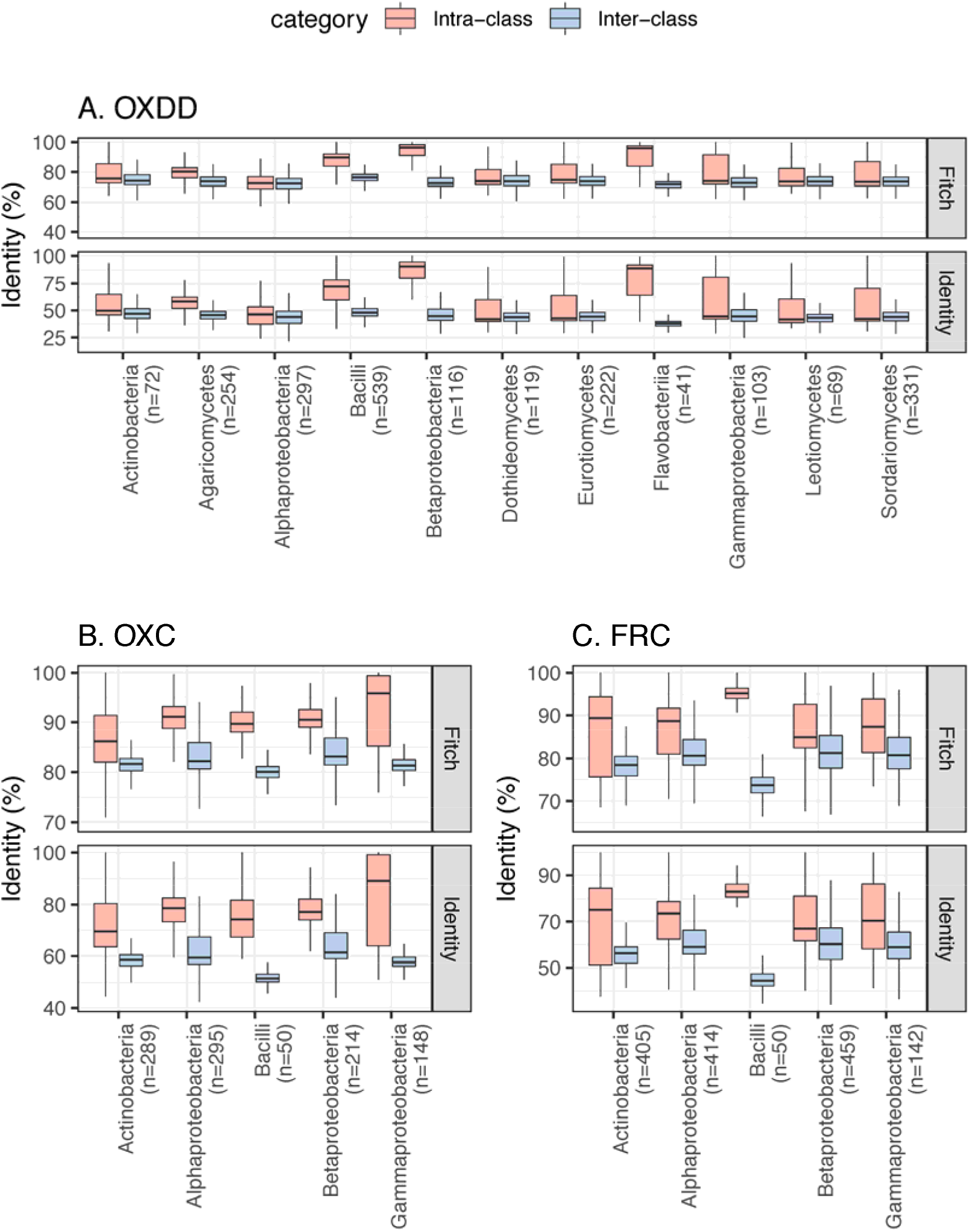
Inter-class and intra-class ODE protein identity associated with each microbial class. Panels focus on OXDD **(A)**, OXC **(B)**, or FRC **(C)**. The pairwise identity between any two protein homologs was calculated based on the multiple alignments using amino acid sequences, by Fitch [105] or identity distance matrix (See Methods for details). The number of ODE homologs available for each class is indicated in parenthesis. Classes with >20 ODE are shown.

**Figure S2.**
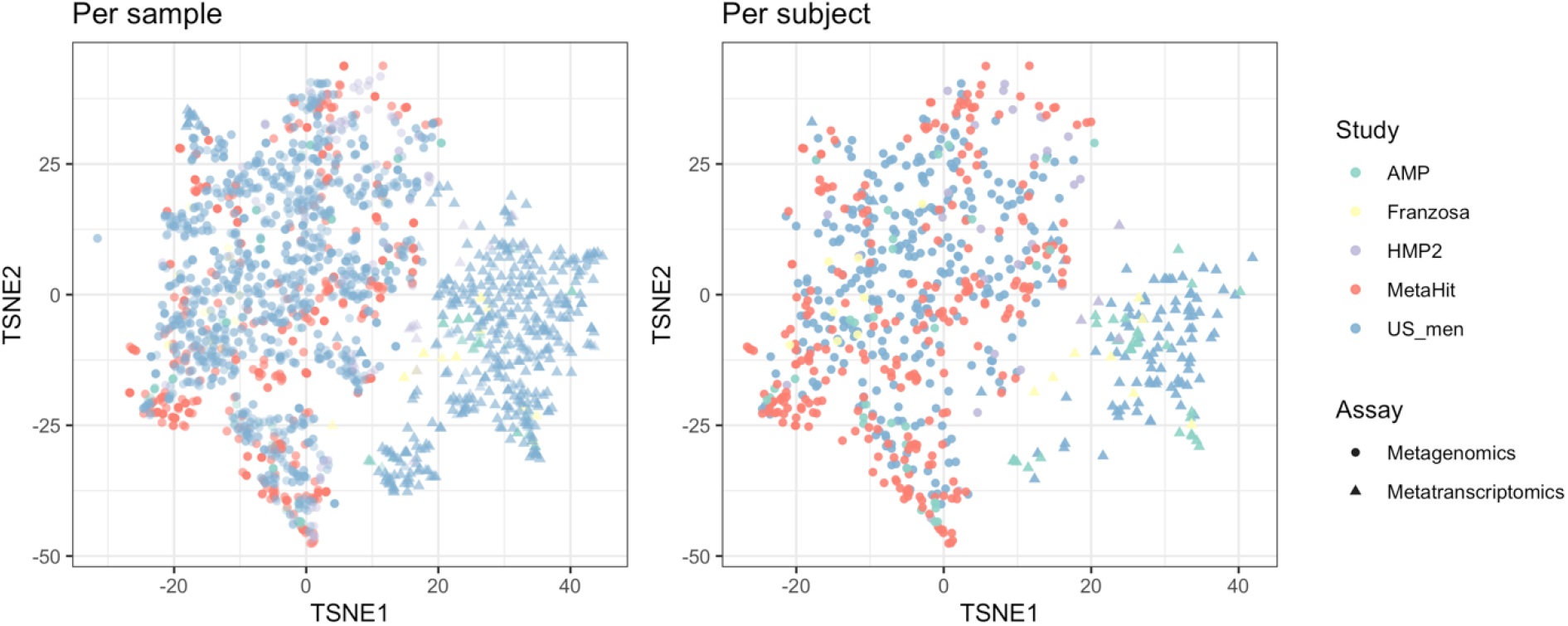
Beta-diversity of metabolic profiles associated with the metagenomic and metatranscriptomic samples from healthy human subjects, ordinated on a Tsne (t-distributed stochastic neighbor embedding) plot. The metabolic profile is assessed by the HUMAnN2 [96] pipeline using (see Methods). The metabolic profiles for each subject are calculated by taking the mean measurements provided. The table shows the number of subjects who provided metagenomic (MTG ▴) and metatranscriptomic (MTS ▲) data. See **Table 1** for study information.

**Figure S3.**
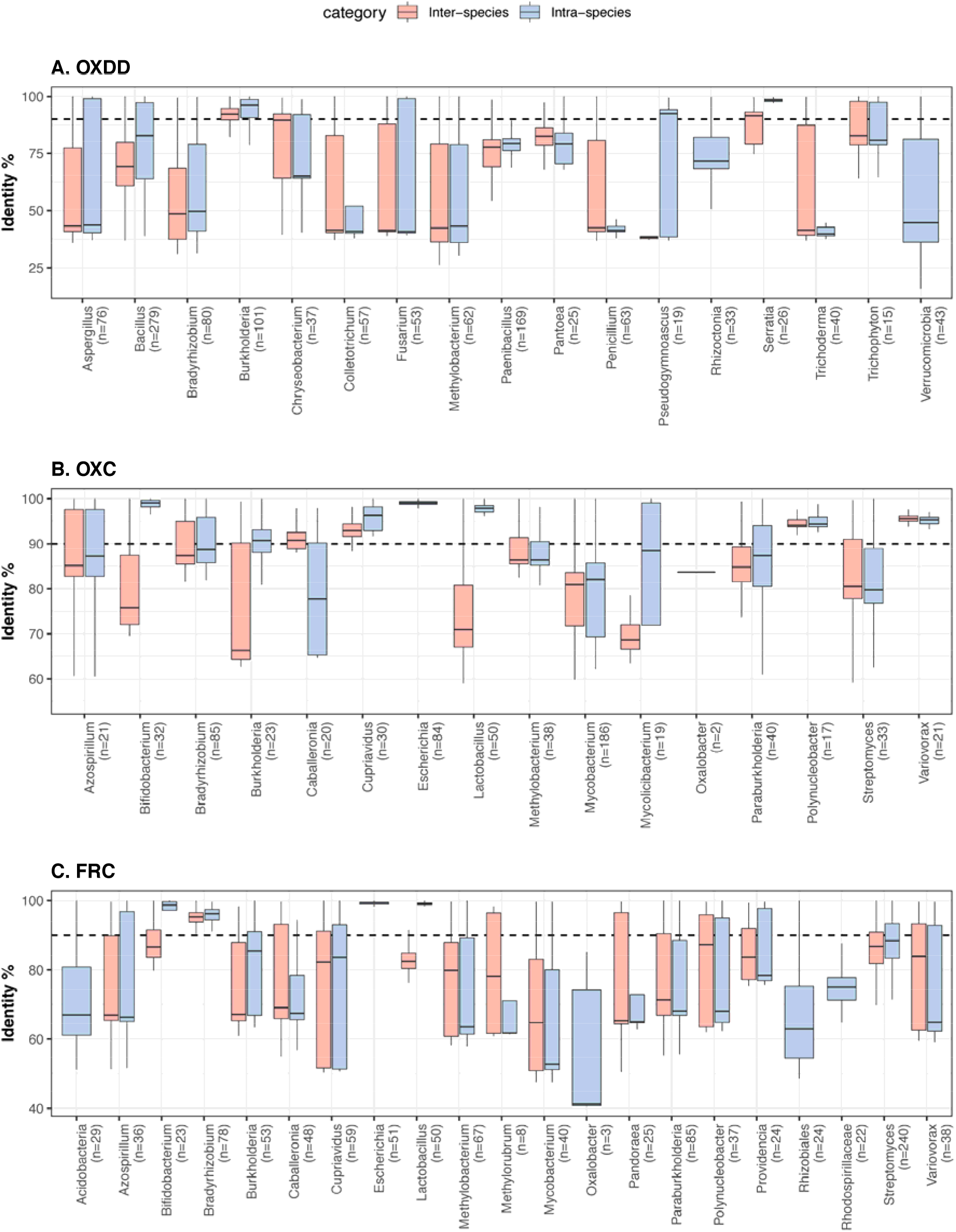
The protein identity between inter-species and intra-species ODEs, for each microbial genus. Panels focus on OXDD(A), OXC (B), or FRC (C). The pairwise protein identities were calculated based on amino acid sequence alignment (See Methods for details). The number of ODE homologs available for each genus is indicated in parenthesis. The blastx identity cutoff 90% used in this study is indicated by the dashed line. Genera with >20 ODE homologs and Genus *Oxalobacter* are shown.

**Figure S4.**
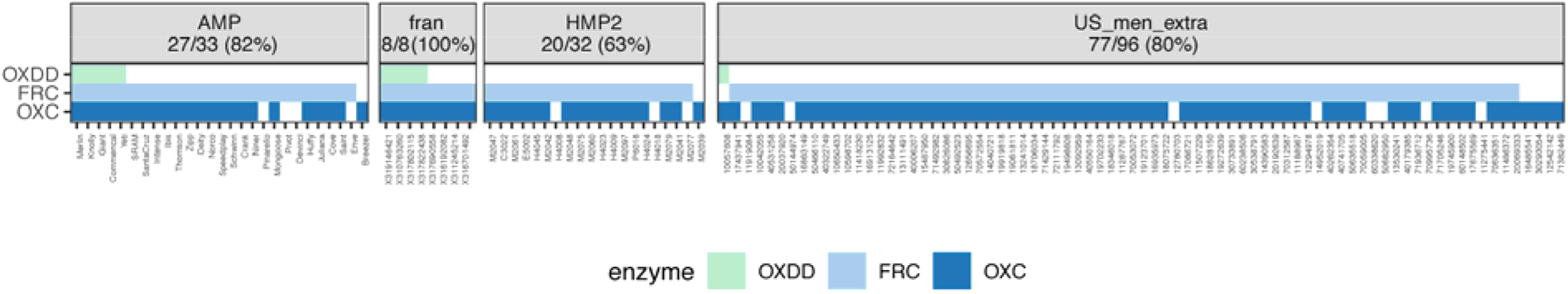
Co-detection of OXDD, FRC, and OXC in the metatranscriptomes of subjects across different studies. Subjects with at least one ODE detected in the metatranscriptome are shown, with percent of total subjects displayed in panels, for each study, indicated in parentheses.

**Figure S5.**
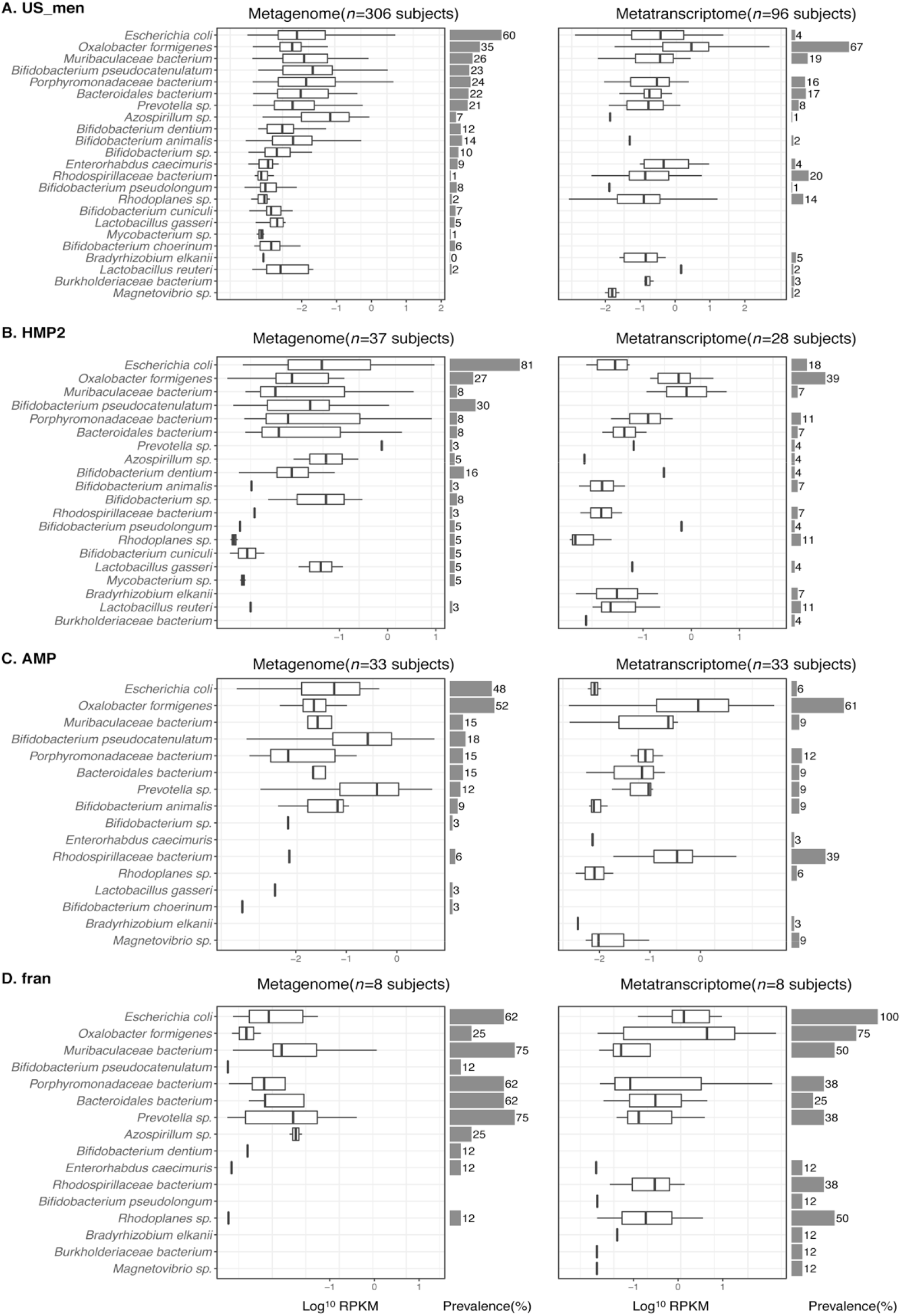
Detection of OXC of microbial species in the microbiome of healthy human subjects from US_men (A), HMP2 (B), AMP (C), or fran (D) study. Left and right panels focus on detection in metagenomic and metatranscriptomic data, respectively. (Follows legend of **Fig. 3A).**

**Figure S6.**
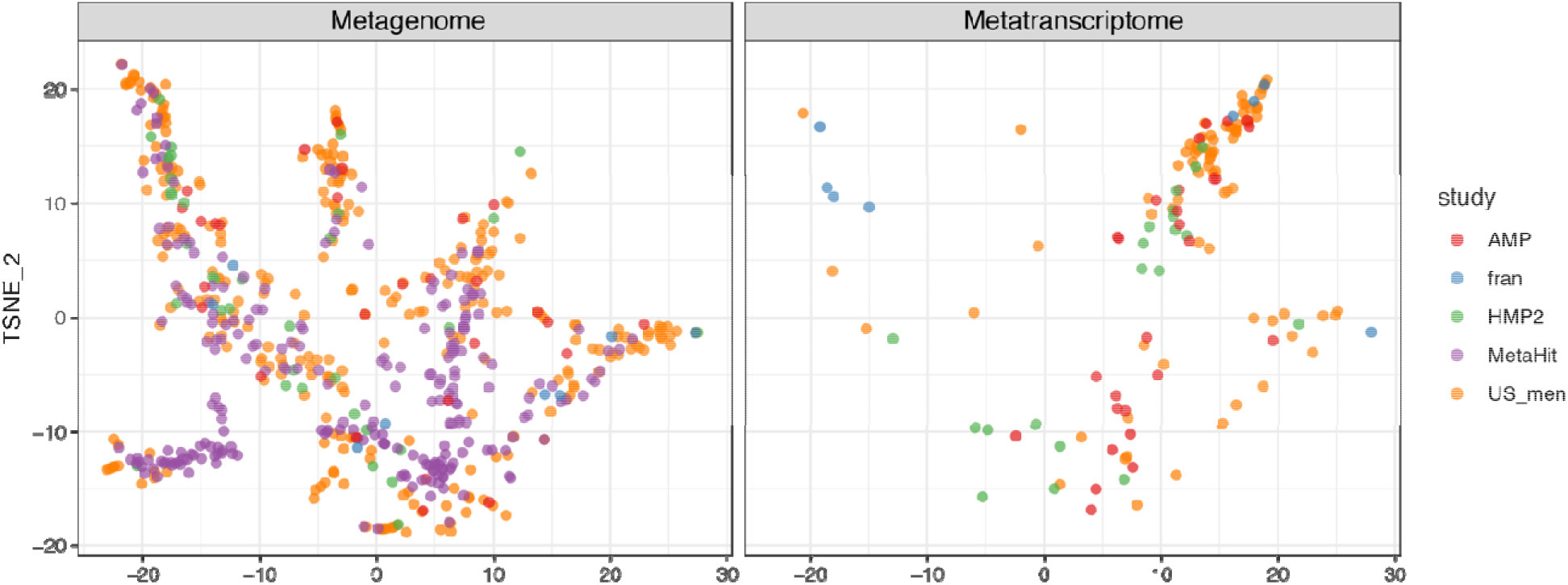
Tsne plot of 594 metagenomic and 131 metatranscriptomic samples, based on the abundances OXC and FRC. OXC and FRC of microbes in **Figures 3A and S5** were used. Tsne is calculated with *Rtsne v0*.*15* package in R. The OXC- and FRC-specific study effects are not significant, examined using PERMANOVA using 1000 permutations (p>0.1).

**Figure S7.**
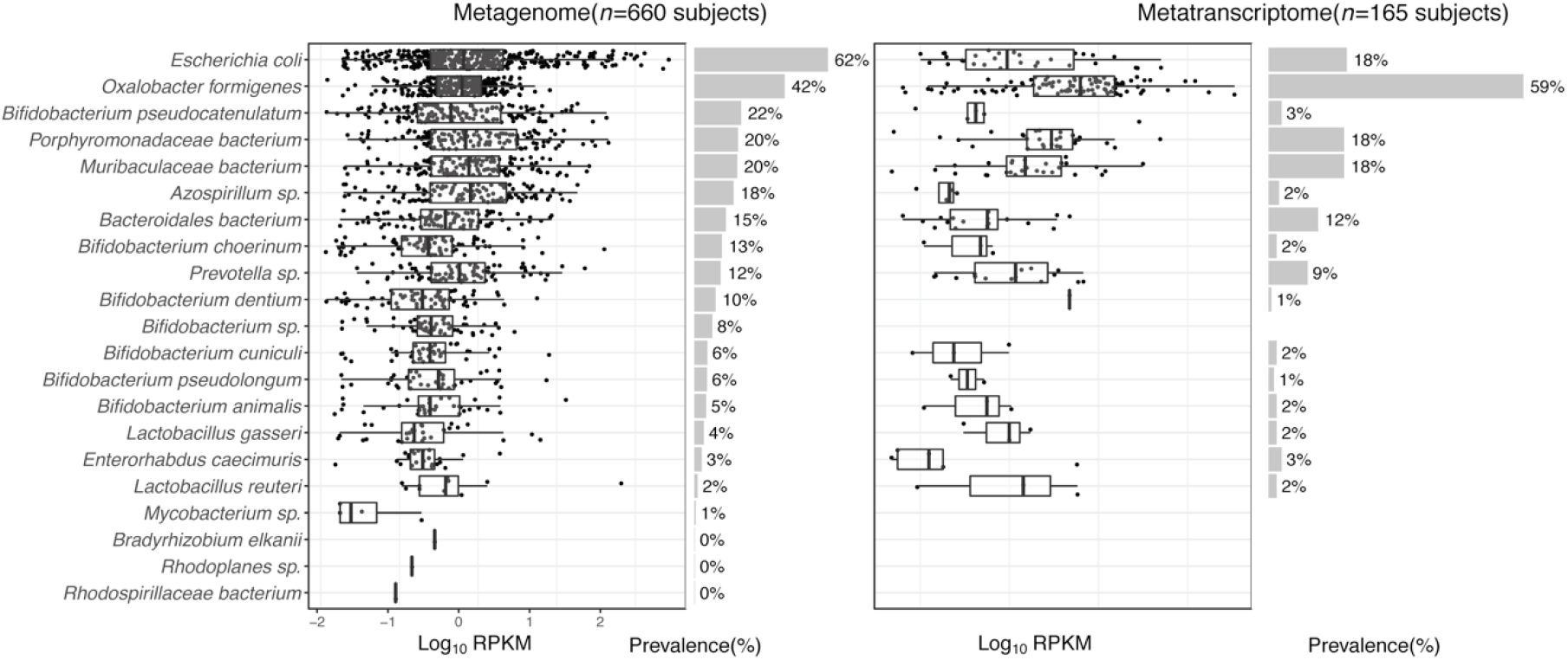
Detection of FRC of microbial species in the metagenome (left) or metatranscriptome (right) of healthy human subjects. (Follows the legend of Fig. 3A).

**Figure S8.**
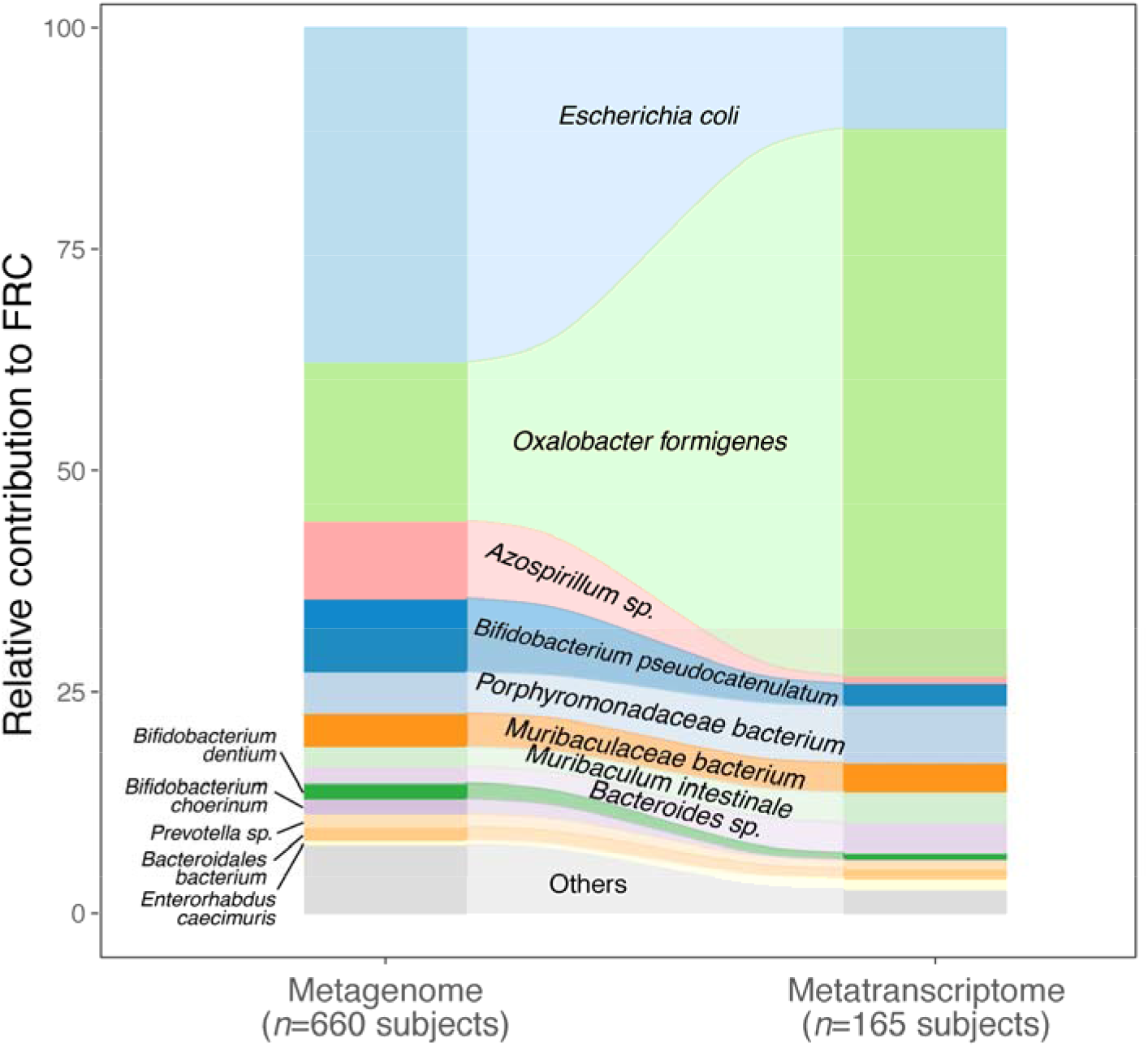
Population-level contribution of individual species to metagenomic (left) or metatranscriptomic (right) FRC. Follows the legend of Fig. 3D.

**Figure S9.**
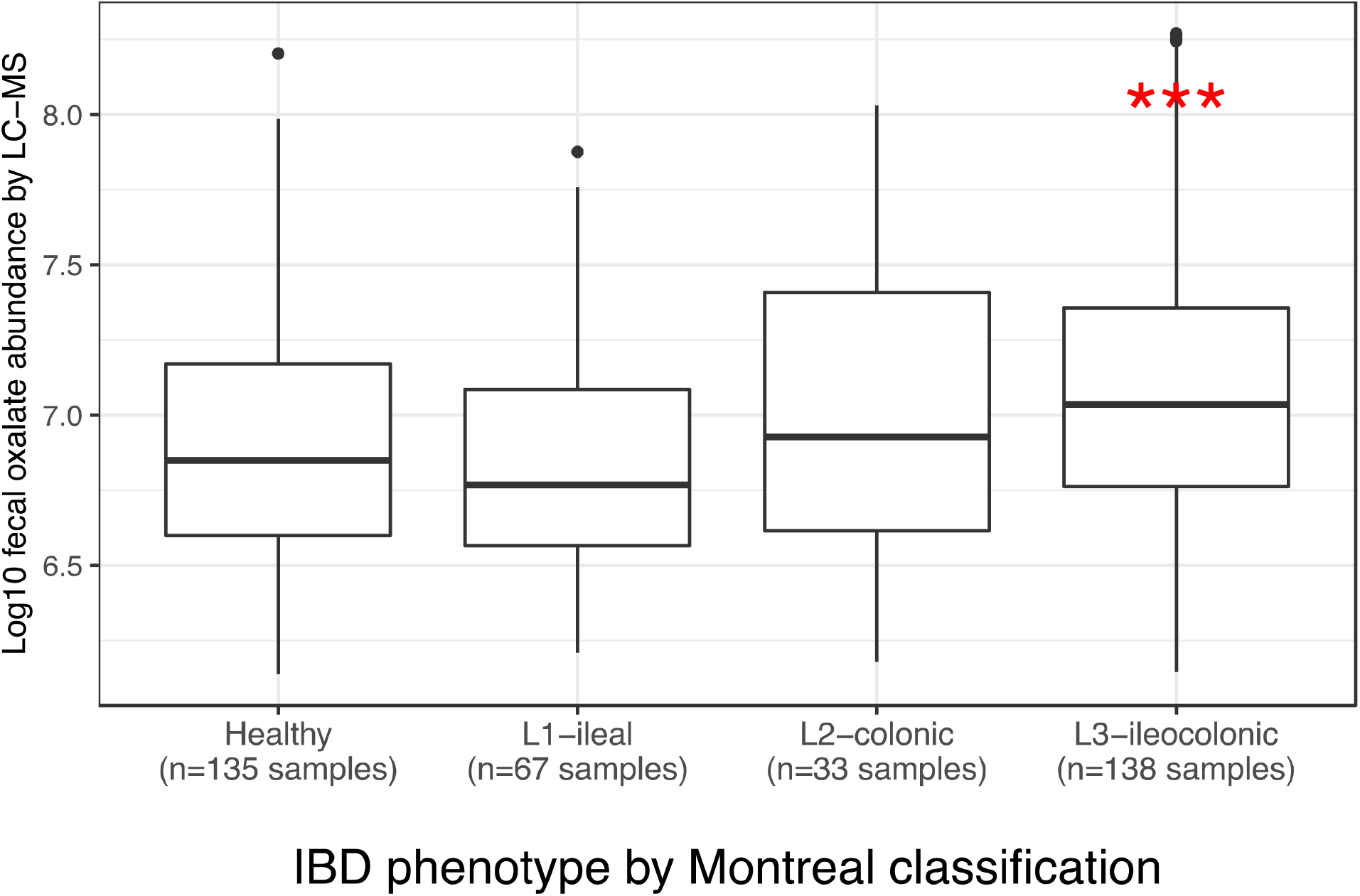
Fecal oxalate concentrations in CD patients, according to the Montreal clinical classification. \*\*\**p* < 0.001 by Mann-Whitney test.

**Figure S10.**
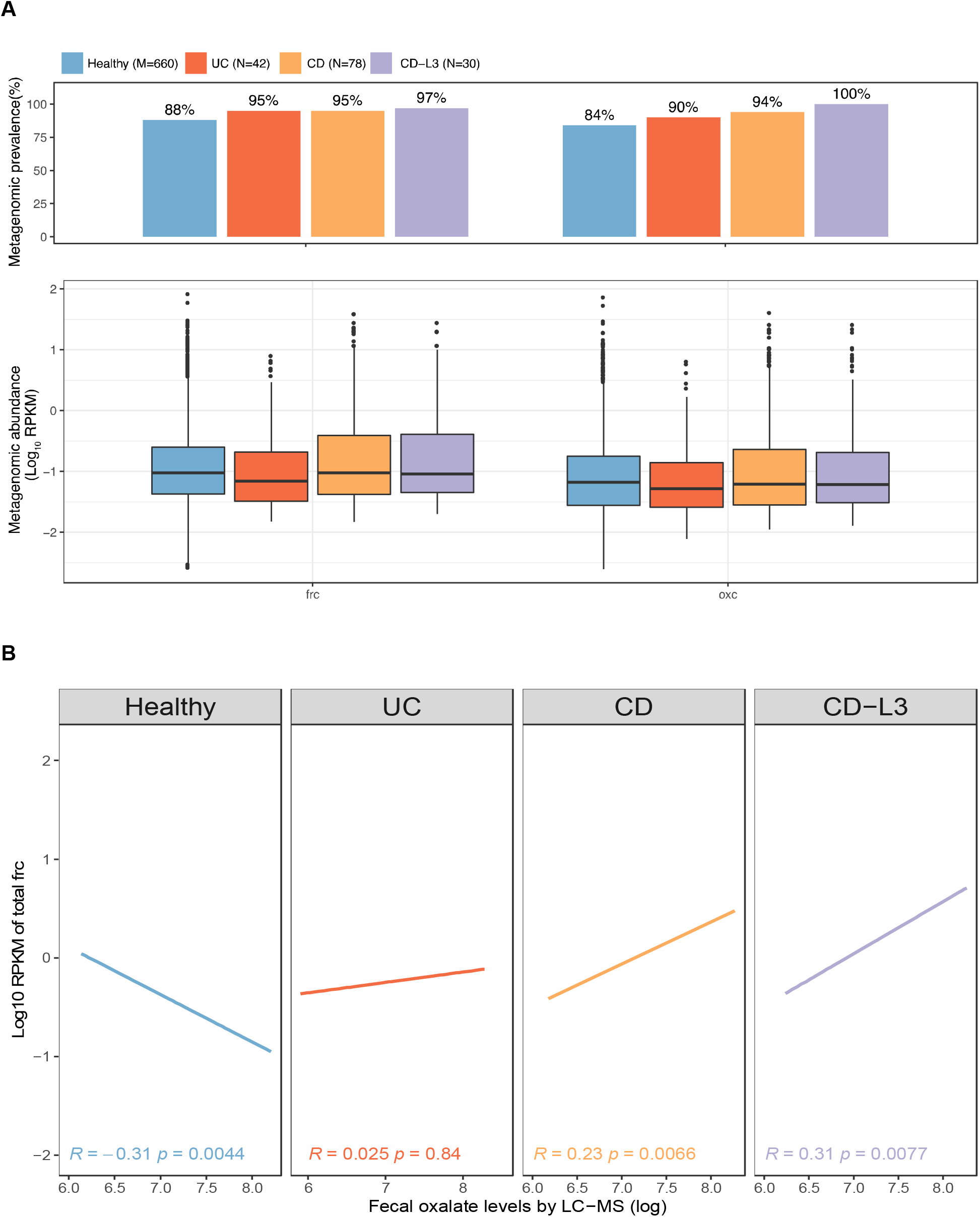
**(A)** Metagenomic prevalence (top) and abundance (bottom) of *frc* and *oxc* in healthy, UC, CD, and CD-L3 subjects. **(B)** Spearman correlation of fecal oxalate and total metagenomic OXC abundance.

